# Are German endoscopy units prepared for the COVID-19 pandemic? A nationwide survey

**DOI:** 10.1101/2020.04.19.20071340

**Authors:** Jakob Garbe, Stephan Eisenmann, Clara S. Heidemann, Marko Damm, Sebastian Krug, Steffen Walter, Frank Lammert, Kaid Darwiche, Patrick Michl, Jonas Rosendahl

**Affiliations:** Department of Internal Medicine I, University Hospital Halle, Germany; Department of Medical Psychology, University Hospital Ulm, Germany; Department of Internal Medicine II, Saarland University Hospital, Germany; For the German Society of Gastroenterology, Digestive and Metabolic Diseases (DGVS); West German Lung Center, Ruhrlandklinik, University Essen-Duisburg, Germany; For the German Society for Pneumology (DGP)

**Keywords:** COVID-19, SARS-CoV-2, endoscopy units, personal protective equipment, endoscopy activity, public health

## Abstract

**Objective:** The COVID-19 pandemic challenges health care systems worldwide. In this situation, guidelines for health care professionals in endoscopy units with increased risk of infection from inhalation of airborne droplets, conjunctival contact and faeces are urgently needed. Recently, the European Society of Gastrointestinal Endoscopy (ESGE) and the German Society for Pneumology (DGP) issued recommendations. However, real-world data on the conditions and requirements of endoscopy units to adhere to this guidance are missing.

**Design:** We conducted an internet-based survey among German endoscopy units from all levels of care from April 1^st^ to 7^th^, 2020. The survey comprised 33 questions and was distributed electronically by the German Society of Gastroenterology, Digestive and Metabolic Diseases (DGVS) and the DGP.

**Results:** In total, 656 endoscopy units completed the survey. Overall, 253 units (39%) cancelled fewer than 40% of their procedures. Of note, private practices cancelled less procedures than hospital-based units. Complete separation of high-risk and COVID-19 positive patients was achieved in only 20% of the units. Procedural measures were well adopted, with 91% of the units systematically identifying patients at risk and 85% using risk-adapted personal protective equipment (PPE). For the future, shortages in PPE (81%), staff (69%) and relevant financial losses (77%) were expected.

**Conclusion:** Concise definitions of non-urgent, elective interventions and endoscopic surveillance strategies are needed to better guide endoscopic activity and intervention cancellations. In the short term, a lack of PPE can constitute considerable impairment of endoscopy units’ operability and patient outcomes.

**SUMMARY BOX:**

1. What is already known about this subject?
  - Recent data indicate a potentially important role of the gastrointestinal tract in the spreading of COVID-19.
  - Endoscopy units and their personnel are at high risk to be exposed to and to distribute COVID-19 infections.
  - Several societies have formulated guidance for endoscopy units in the current situation, but their feasibility is unclear.
2. What are the new findings?
  - Endoscopic activity seems not to be limited to urgent interventions across all units as 39% of all endoscopy units cancelled less than 40% of procedures.
  - For most endoscopy units, structural conditions are insufficient to realize a complete separation of high-risk patients, which can be guaranteed by only 20% of the units.
  - The willingness to adhere to the recommendations is very high, as most endoscopy units adopted their procedures accordingly. Shortage of personal protective equipment is a critical concern in many units.
3. How might it impact on clinical practice in the foreseeable future?
  - An update of the current recommendations to refine practicable measures for the majority of endoscopic units is warranted.
  - A concise definition of non-urgent or elective procedures as well as postponement strategies and intervals are of utmost importance, since current data implicate that transmission of SARS-CoV-2 via the respiratory and gastrointestinal tract may be critical for public health.

## INTRODUCTION

The on-going Coronavirus disease 2019 (COVID-19) pandemic is a challenge for patients, healthcare professionals and populations worldwide. Endoscopy units and their staff are at high risk to be exposed to and distribute the severe acute respiratory syndrome coronavirus 2 (SARS-CoV-2). Therefore, guidance on endoscopic activity in this pandemic is urgently needed. Recommendations have to balance safety issues and practicability and therefore need to be crossvalidated by real-world data and updated as new findings emerge.

Endoscopic procedures, especially those of the upper gastrointestinal (GI) tract and the upper as well as lower airways, are associated with increased formation of potentially infectious aerosols. Consequently, the risk of infection in endoscopy units is significantly increased [1]. Recently, evidence emerged that live SARS-CoV-2 is excreted in the faeces, indicating a potential risk of infection also during lower GI endoscopy [2–4]. So far, no endoscopy-associated COVID-19 infections of health care workers or patients have been reported. However, an estimated infection rate of 20% of health care workers during the north Italian outbreak highlighted the eminent risk for medical personnel [5].

To address the need for guidance in this situation, the European Society for Gastrointestinal Endoscopy (ESGE) and the German Society for Pneumology (DGP) have published recommendations for endoscopy units on structural and procedural aspects of patient and equipment handling to mitigate infection risks [1, 6]. To date it is unclear whether these recommendations can be implemented in the majority of endoscopy units as data are scarce and limited to few centres [7]. We conducted this survey to explore the ability to implement these recommendations and learn about different approaches across different centres, both hospital-based and private practice-based. As such our data provides a basic framework for future recommendations.

## METHODS

Ethical approval was obtained from the local ethical review committee (Title: “Versorgungsrealität in deutschen Endoskopiezentren in Zeiten der COVID-19-Pandemie”; 2020-044; March 30^th^, 2020). Based on the ESGE and DGP recommendations, we developed 33 questions to assess the adherence to and practicability of the guidelines in an internet-based survey. Relevant questions were developed to cover structure-related, personnel-related and procedure-related measures. Where appropriate, we added additional items to better understand the local situation.

In a fourth category, expectations for the future of endoscopy units were investigated. Answers for questions on measures were yes/no-questions or multiple choice, few allowed for answers or specifications in free text. Expectations for the future were requested on a 5-step Likert scale (highly probable to highly improbable). The survey was constructed following expert recommendations of S.W. It was addressed to the heads of endoscopy units that were in addition asked to provide basic information on the characteristics and care levels of their units. The full and translated survey is provided in the supplements.

The software LimeSurvey (LimeSurvey GmbH, Germany) was used to conduct the survey online. The survey was distributed via the German Society of Gastroenterology, Digestive and Metabolic Diseases (DGVS) and the German Society for Pneumology (DGP). Completion of the survey was possible for 7 days from April 1^st^ to April 7^th^. Descriptive statistics were calculated using Office Excel 2016 (Microsoft Corporation, Redmond, US).

## PATIENT AND PUBLIC INVOLVEMENT

Patients and the public were not involved directly in the current study.

## RESULTS

Overall, 676 complete questionnaires were retrieved, of which 15 (10 from private practices and 5 from hospitals) were identified as duplicates and not included in the analysis. Duplicates were identified using the provided postal code and answer patterns as well as cross-referencing in the databases for hospitals, practices and outpatient clinics [8–10]. Three questionnaires from Switzerland and two from Austria were also excluded. Of the remaining 656 questionnaires, 393 endoscopy units were from hospitals and 263 from outpatient clinics and practices all over Germany (**Fig. 1**). In total, 145 (22.1%) of the responses did not include the postal code. In the timeline of the pandemic, the survey fell into a phase of continued but slowed exponential growth (**Fig. 2**).

**Figure 1:**
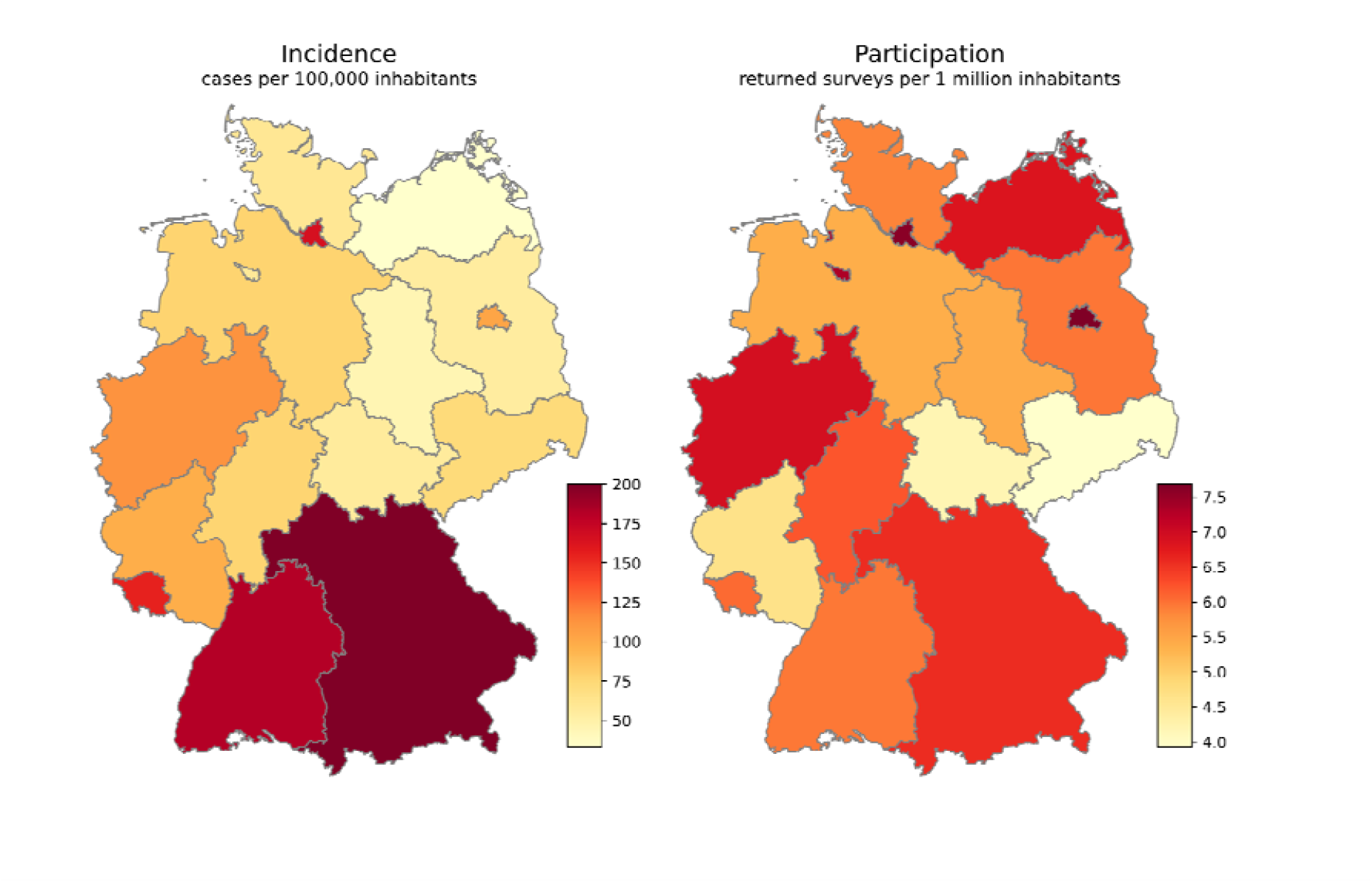
Geographical heatmap of COVID-19 incidence rates in federal states on April 7, 2020 and participating endoscopy units. Nationwide incidence was 119/100,000 inhabitants on April 7. Of note, 145 anonymous responses (22.1% of all responses) are not plotted. COVID-19 data were retrieved from the Robert-Koch-Institute, Germany [19].

**Figure 2:**
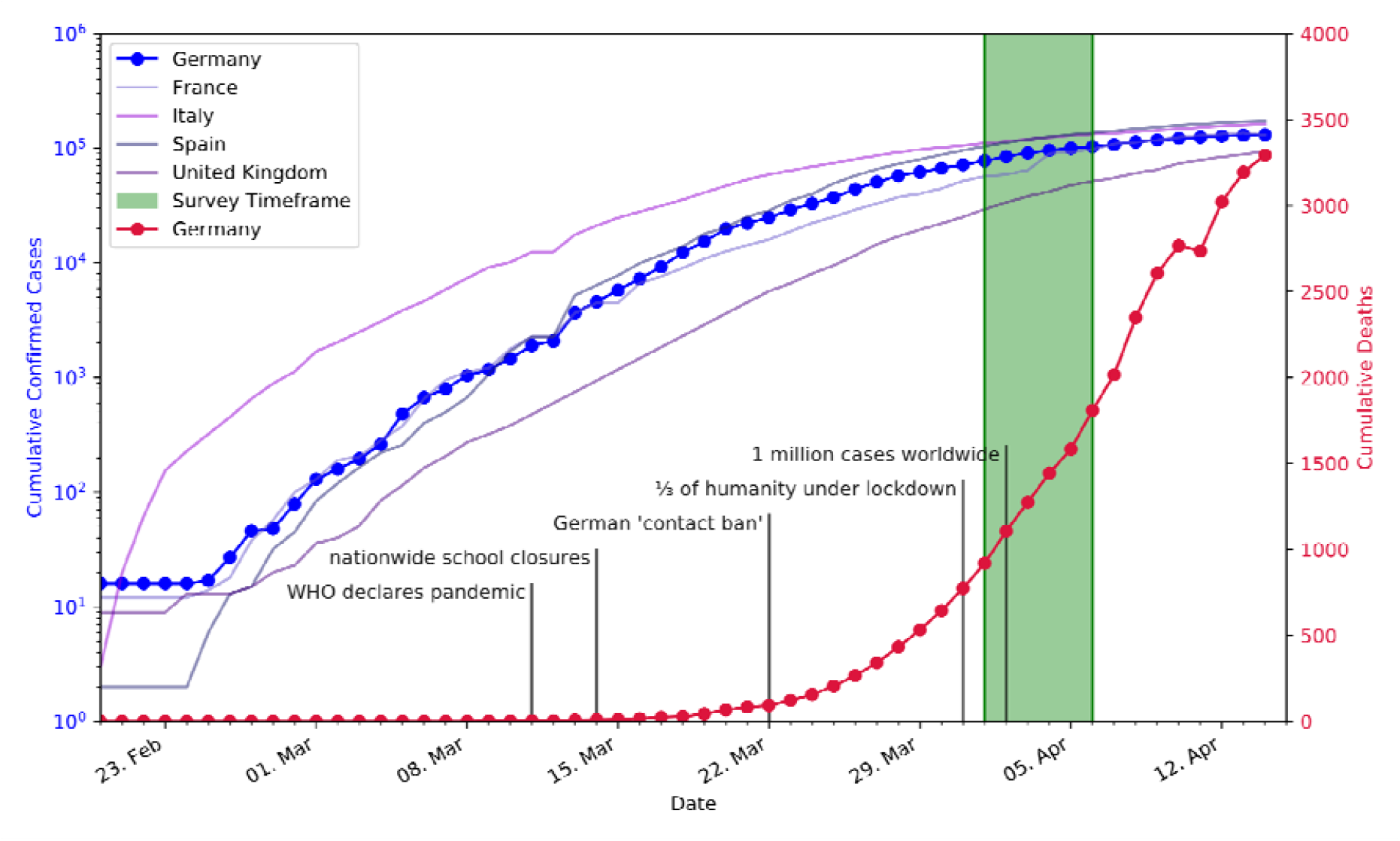
Survey timeframe in context of recent major events and progress of the COVID-19 pandemic. Neighbouring European countries are plotted for comparison. For clarity reasons, cumulative cases are plotted on a logarithmic scale. Data were retrieved from the Johns Hopkins University, US [20].

**Table 1** shows selected survey results. A comprehensive documentation of the survey results can be found in the **Supplementary Tables 1 – 4**.

**Table 1:**
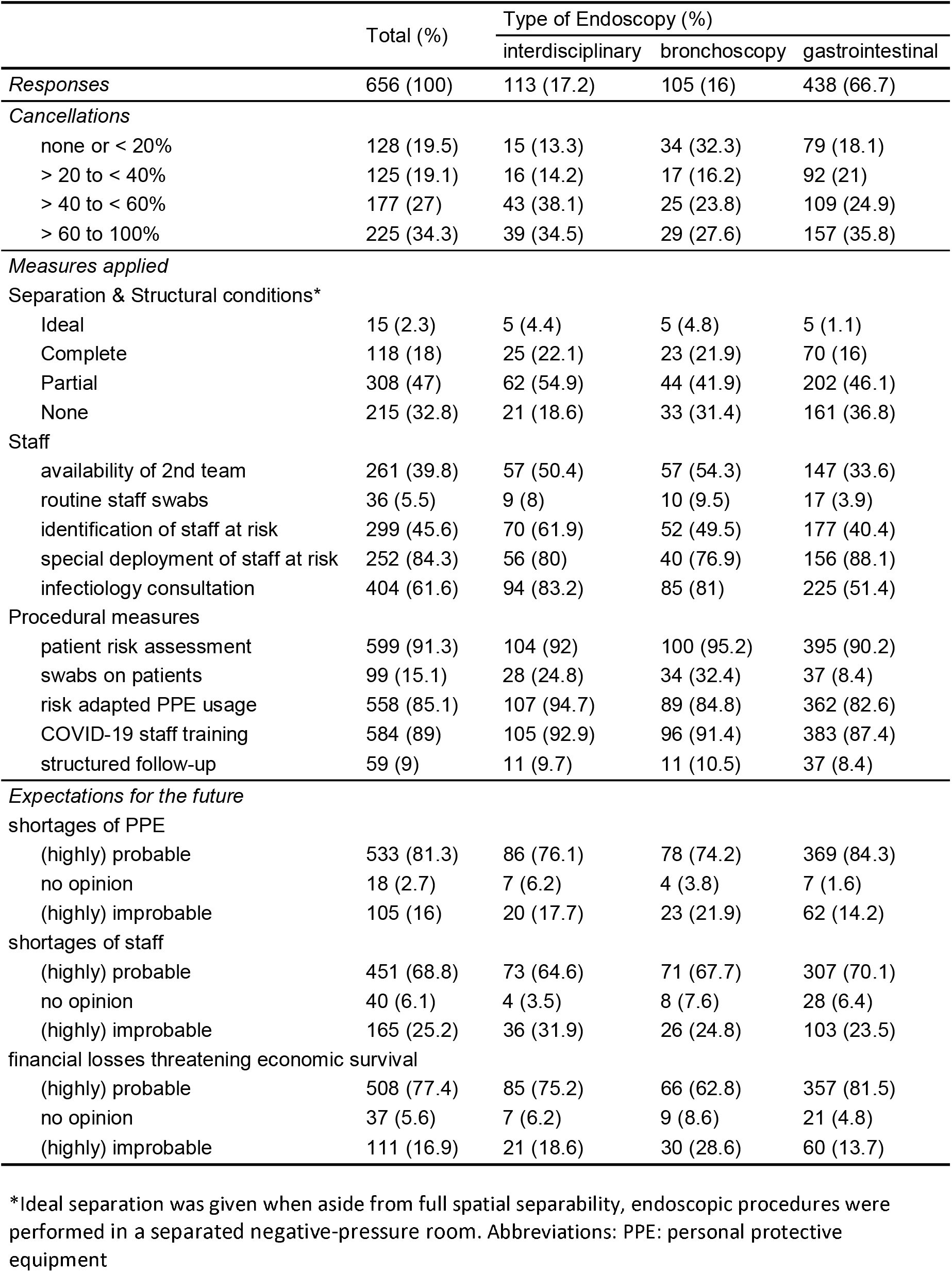
Condensed survey results by type of endoscopy unit.

### Cancellations of interventions

Overall, 253 endoscopy units (38.6%) cancelled less than 40%, whereas 225 (34.3%) cancelled more than 60% of their procedures. Interestingly, 45.6% of private practices cancelled less than 40% of their procedures as compared to 33.8% of hospital-based units (**Supplementary Table 3**). In the high prevalence Southern and Western regions of Germany, a larger fraction of endoscopies was cancelled (37.8%) than in the low prevalence areas in Northern and Eastern Germany (26.5%) (**Supplementary Table 4**).

### Separation of patients at risk

Ideal conditions for separation of infectious patients were achieved in 2.3% of endoscopies only (n = 15), where a complete spatial separation and a negative pressure endoscopy suite were available. A complete separation without negative pressure suites in the unit or ideal separation was possible in 20.3% of the investigated endoscopy units. In general, most units were not capable to separate well, as 47% could only separate partially (periinterventional area or endoscopy room) and 32.8% could not separate high-risk or proven COVID-19 patients at all.

### Staff

For many endoscopy units there was some capability to compensate shortages in personnel: 40% of endoscopies had a second “reserve” team in case one team had to move into quarantine. To protect staff, about half of the endoscopy units (45.7%) identified staff members with an elevated risk for a severe course of a COVID-19 infection. In most cases (84.3%), such personnel were deployed in low-risk areas to minimize infection risk.

To define proceedings in the endoscopy units, a hygiene specialist or infectiologist was consulted in 61.6%. Private practices and outpatient clinics consulted these specialists to prepare for the pandemic less frequently (24.8%) compared to hospital-based endoscopy units (86.5%) (**Supplementary Table 3**).

### Procedural measures

Most endoscopy units (91.6%) identified patients at risk of infection in a structured manner. Here, Bronchoscopy units seemed to be more rigorous in comparison to Gastroenterological units (95.2 vs 90.2%). To mitigate risk of infection, most endoscopy units issued new instructions on the risk adapted personal protective equipment (PPE) use and trained their staff in the handling of COVID-19 patients (>85%).

### Swabs

Routine swabs for personnel were performed in 5.5% of endoscopy units. Also, the overall rate of pre-interventional swabs for patients was low (15.1%). Bronchoscopy units had a considerably higher rate of pre-interventional swabs (32.4%) compared to GI units (8.4%).

### Expectations

The majority of endoscopy units (81.3%) perceived shortages in PPE as probable or highly probable. PPE shortages were especially expected by private-practices and outpatient clinics (92.4%) (**Supplementary Table 3**). Overall, 68.8% of endoscopy units expected staff shortages during the pandemic. In total, 77.4% of endoscopy units anticipated substantial financial losses that could threaten the economic survival to be probable or highly probable. In GI units this expectation was even worse (over 80%), compared to Bronchoscopy units (61%).

## DISCUSSION

This is the first nationwide survey to obtain real-world data on how endoscopy units cope with the current COVID-19 pandemic and the measures taken to ensure a continued balanced patient care. The framework of this survey relied on the ESGE and DGP recommendations that had been published on March 18^th^, 2020, 14 days before the current survey was conducted [1, 6]. The ESGE guidance statement was supported by the German Society of Gastroenterology, Digestive and Metabolic Diseases (DGVS) and disseminated to all members via email on March 19, 2020.

### Cancellations and surveillance endoscopies

In recent reports the prevalence of GI symptoms in SARS-CoV-infected patients was higher than previously estimated ranging from 11.4 to 61.1% in two series with a special focus on GI symptoms [11, 12]. Moreover, there is emerging evidence that SARS-CoV-2 is excreted in the faeces [2], even after it becomes undetectable in the pharynx and lung [13]. Furthermore, live virus was detected in the stool of patients and therefore the faecal route of infection could be relevant for the distribution of COVID-19 [3, 4]. A recent report confirmed independent virus replication in the intestine and virus may be shed in stool even weeks after resolution of symptoms [14]. As a consequence of the current knowledge, endoscopy societies have recommended to postpone or even to cancel all but emergency and absolutely essential endoscopic interventions [1, 15, 16]. Our results demonstrated that approximately one third of the participating endoscopy units most likely followed these recommendations as more than 60% of all procedures were cancelled. However, approximately 40% of the endoscopy units still performed more than 60% of their procedures. It seems unlikely that these 60% of the procedures are all emergency procedures. Interestingly, units performing >60% of procedures were more likely to be private practices. In their comments, heads of out-patient GI units repeatedly pointed out the ethical dilemma since their economic survival is threatened by cancellations. Therefore, strategies to substitute for financial losses endangering economic survival need to be discussed and implemented particularly for privately owned endoscopy units. In addition to financial reimbursement, it seems to be necessary to refine the definitions of non-urgent, elective procedures.

Of note, the British Society of Gastroenterology has published a list of specific interventions and indications in categories of “urgent/emergency”, “case-by-case decision” and “defer until further notice” to guide planning and cancellations, which might serve as a model in this context [16]. As long as sound evidence for the safety of upper and lower GI endoscopy is missing, all endoscopy units should be urged to adhere to the current recommendations and postpone elective endoscopies and surveillance programs.

### Structural and procedural measures

It is important to notice that even in a highly developed health care system like Germany, only 20.3% of all endoscopy units had the capability to conduct interventions of high-risk patients in a spatially separated endoscopy ward. Ideal conditions with a negative pressure endoscopy room were an exception. As pandemics are irregularly recurring events these observations should be taken in consideration when new endoscopy units are planned in the future and when emergency plans are updated.

Furthermore, this limited separation also restricted the ability of unit heads to introduce procedural measures such as employment of staff at risk of severe courses in a low-risk area in the endoscopy ward. Currently, there is no solid evidence that these measures are effective, but the considerable psychological burden for the staff at risk might have been an important factor for heads of endoscopy units to implement such measures. Where identified, a majority of the staff at risk was deployed in areas without patient contact or in low-risk areas. Other procedural measures were implemented well, such as risk stratification of patients, risk adapted use of PPE and training of staff to handle suspected or proven COVID-19 patients. In contrast, routine follow-up to inquire about symptoms and swabs on patients was rarely conducted.

### Shortages in PPE

According to the survey, fear is evident among many endoscopists that a lack of PPE might further complicate endoscopic activity in the future and an overt lack of PPE has substantially impacted private practices already. Several measures have been undertaken to overcome this shortage. The German Society for Sterile Supply has published recommendations on the reprocessing of masks, which is possible to a limited extent [17]. While many practices and outpatient clinics do not have the necessary tools to provide safe reprocessing, cooperation with nearby hospitals and laboratories might be an option to reduce the impact of current PPE scarcity.

### Strengths & limitations

Since there are no absolute numbers for endoscopic units in Germany available, we cannot calculate the exact response rate of our survey. For GI units in hospitals a rough estimate can be made using intervention numbers from responses and published GI intervention numbers of inpatients [18]. With this data we estimated that between 30 to 50% of hospitals with a GI endoscopy unit answered our survey. Furthermore, our responses are well distributed throughout Germany indicating representativeness. Lastly, the short survey timeframe allowed for a concise snapshot of the situation in a phase of rising case numbers and uncertainty.

Our study was limited to Germany as the COVID-19 pandemic was in a relatively uniform state during the survey. Otherwise, this limitation implies that the transferability of some results to other health care systems and countries may not be possible. Finally, the survey offered participants the possibility to remain anonymous. Where postal codes were given, we were able to discard duplicates, however, as 22.1% of the responders did not provide their postal codes a certain bias cannot be excluded with certainty.

## CONCLUSION

Procedural measures from ESGE and DGP recommendations were broadly adopted and can be implemented easier than structural measures for which ad hoc realization is naturally limited.

In the short term, the shortage of PPE might be the most important obstacle in maintaining a safe endoscopy environment. Cooperation with other health care facilities to safely reprocess PPE might be a way to maintain operability of outpatient clinics and practices.

Fractions of cancelled interventions differed considerably and were distributed along the spectrum. Considering rising evidence of vital SARS-CoV-2 in faeces, the continuation of surveillance colonoscopies and other elective endoscopies might constitute a health risk and affect patient outcomes. On a political level, societies, associations and health authorities should consent on the definition of non-urgent, elective endoscopies.

## Data Availability

Deidentified aggregate parcipant data are available upon reasonable request to the corresponding author.

## AUTHOR CONTRIBUTIONS

J.G., S.E. and J.R conceived, designed and directed the study.

J.G., S.E., and J.R drafted and revised the manuscript with substantial help from P.M.

J.G., C.H. and M.D. performed statistical work and contentual review of survey responses.

S.W. guided the survey conception.

All other co-authors helped to develop and disseminate the survey. All authors approved the final manuscript and contributed critical revisions to its intellectual content.

## COMPETING FINANCIAL INTERESTS

The authors declare no competing financial interests.

## FUNDING

The authors have not declared a specific grant for this research from any funding agency in the public, commercial or non-profit sector.

## ACKNOWLEDGEMENT

The authors thank all participating heads of endoscopy units for their responses, trust and insightful comments.

## Abbreviations

COVID-19: Coronavirus disease 2019
DGP: German Society for Pneumology
DGVS: German Society of Gastroenterology, Digestive and Metabolic Diseases
ESGE: European Society for Gastrointestinal Endoscopy
GI: gastrointestinal
PPE: Personal protective equipment
SARS-CoV-2: severe acute respiratory syndrome coronavirus 2

## Notes

### Competing Interest Statement

The authors have declared no competing interest.

### Funding Statement

There is no funding to be reported in connections with this publication.

